# Incident Stroke in Pediatric Sickle cell Anemia Despite Overall Improved Transcranial Doppler Velocity in a Ugandan Hydroxyurea Trial: Antecedent and ongoing risks

**DOI:** 10.1101/2025.01.28.25320389

**Authors:** Bill Wambaka, Amiirah Mpungu, Vincent Mboizi, Dennis Kalibbala, Grace Nambatya, Susan Murungi, Maxencia Kabatabaazi, Maria Nakafeero, Phillip Kasirye, Deogratias Munube, Ruth Namazzi, Richard Idro, Nancy S. Green

**Affiliations:** Global Health Uganda, Kampala, Uganda; Division of Hematology, Cincinnati Children’s Hospital Medical Center, Cincinnati, Ohio, USA; Dept. of Paediatrics and Child Health, Makerere University School of Medicine, Kampala, Uganda; Division of Hematology, Oncology and Stem Cell Transplantation, Dept. of Pediatrics, Columbia University Medical Center, New York, NY, USA

**Keywords:** Sickle cell anemia, sickle cell disease, stroke, hydroxyurea, clinical trial

## Abstract

**Introduction:** Transcranial doppler ultrasound (TCD) screening for primary stroke prevention in children with sickle cell anemia (SCA) was established in higher-resource regions, targeting interventions for highest velocity (“abnormal”). We sought to identify additional stroke risk factors in Uganda.

**Methods:** We conducted a 30-month open-label single-arm Ugandan hydroxyurea trial, dose-escalated to maximum tolerated dose, aimed to test brain protection for children aged 3-9 years with SCA. Study procedures included history, clinical stroke examination and prospective TCD and laboratory assessments.

**Results:** Overall, 264 children received study HU, mean age 5.6±1.7, hemoglobin 7.8±1.2g/dL, fetal hemoglobin (HbF) 11.9±8.1%, enrolment TCD maximum velocity 148.4±29.3cm/second; 15 (5.7%) had abnormal TCD. Mean TAMV at trial completion was131.9±SD25.7 cm/sec. Four participants without abnormal enrolment TCD developed acute stroke within the initial 16 months (incidence 0.62 per 100 person years); Two had enrolment HbF ≤3.1%, 2 had low oxygen saturation (90%), 1 had recurring severe anemia necessitating multiple transfusions. Apparent stroke precipitants were severe malaria, acute chest syndrome, recent pain crisis or uncertain cause. At trial completion, 8 additional participants had a higher risk TCD category than at enrolment.

**Conclusion:** Effectiveness of TCD screening for stroke prevention may vary by region, as no participant with incident stroke was at highest risk. Antecedent and/or ongoing SCA-related risks of anemia, low HbF, hypoxemia, infections and/or disease complications likely contributed to stroke despite trial HU. Results suggest that TCD alone may not fully identify highest stroke risk in the region, and need for primary stroke prevention from early and continuous hydroxyurea therapy.

## Introduction

Approximately 7.7 million people worldwide live with sickle cell disease (SCD), the majority in sub-Saharan Africa (SSA).^1^ Sickle cell anemia (SCA), generally a more severe phenotype for cerebrovascular injury than the other genotypes, increases the risk of stroke by over 200-fold compared to unaffected children.^2,3^ High risk of stroke leads to considerable morbidity and mortality in this population. In higher-resource settings, transcranial doppler ultrasound (TCD) screening for abnormal cerebral artery blood flow velocity is the established strategy for identifying the highest at-risk children for primary stroke and instituting preventative therapy.^4,5^ This strategy uses standard risk categories of time-averaged maximum mean velocity (TAMV) that were established in a U.S. pediatric longitudinal cohort in the landmark STOP Trial.^6^ Screening results are deemed “abnormal” for TAMV ≥200cm/second in any of the major insonated vessels.^2^ These same criteria are used globally for pediatric SCA stroke risk stratification and intervention of risk reduction.^7-10^ Hydroxyurea (HU), the most widely used and effective disease-modifying medication for children with SCA, is recognized as reducing stroke risk by lowering TAMV.^2,11^ HU is the treatment of choice for risk reduction in parts of SSA, when safe blood transfusion therapy may not be regularly available.^11-14^

Using the same TCD criteria, our prior cross-sectional study of children with SCA in Uganda, “Burden and Risk of Neurological and Cognitive Impairment in Pediatric Sickle Cell Anemia in Uganda (BRAIN SAFE)” demonstrated evidence of higher stroke risk by standard TCD screening, with 2.0% having an “abnormal” and 15.1% having “conditional” TCD.^15^ The latter category is for intermediately elevated TCD TAMV.^2^ However, while TCD screening remains an invaluable tool for stroke risk screening, some children with SCA develop acute stroke even with normal TCD or with known cerebrovascular injury despite chronic transfusion therapy.^9,16^ Clinically unpredictable stroke may be more common in SSA where affected children are exposed to a high burden of additional potential risk factors or precipitants including malarial and other severe infections, malnutrition and undiagnosed hypertension.^14,17^

Our recently completed a 30-month HU treatment trial in Uganda, “BRAIN SAFE-II,” aimed to prevent incident stroke and preserve neurocognitive function in children with SCA aged 3-9 years.^18^ HU was dosed to maximum tolerated dosing to optimize protective effects.^19,20^ Primary outcomes were deaths, clinically detected acute stroke and abnormal TCD TAMV at month 30 compared to enrolment. Baseline trial assessments included standard TCD screening to determine stroke risk. Of the four trial participants with acute stroke during the trial, none had an abnormal TCD at enrolment. In addition, other participants were found to have higher TCD risk category at trial completion than at enrolment. We hypothesized that children with SCA in sub-Saharan Africa have additional identifiable risk factors or precipitants for acute stroke beyond abnormal or elevated TCD despite effective HU therapy.^21^ This study aimed to identify precipitants or risk factors that may predispose children with SCA to acute stroke in the region. Identification of such factors, if amenable to prevention or intervention, may lead to reduced SCA cerebrovascular complications.

## Methods

This was a prospective study on acute stroke in children with SCA nested within our HU trial for prevention of SCA-related brain injury. All study procedures were approved by the Makerere University School of Medicine Ethics Committee, the institutional review boards (IRB) of Columbia University and Mulago Hospital.

Participants were children with SCA who were attending the Mulago Hospital Sickle Cell Clinic (MHSSC) in Kampala.^15^ Children treated there receive care following Ugandan SCA guidelines, including monthly folic acid, malaria prophylaxis and penicillin prophylaxis until the age of 5 years.^18,22^

Trial eligibility required SCA confirmation by hemoglobin (Hb) electrophoresis.^18^ Additional criteria included a total of <6 months of prior HU therapy; no evidence of stroke by a standardized pediatric stroke-focused examination, the pediatric NIH stroke scale (PedNIHSS); not enrolled in another concurrent trial.^15^ Children with a recent clinical event characterized by an acute febrile illness or vaso-occlusive crisis within the preceding 2 weeks, or blood transfusion within 90 days were considered temporarily ineligible until meeting the additional clinical criteria of duration of wellness and or post-transfusion. Legal caregivers of children who met the eligibility criteria from provided written consent, with assent from children ≥8 years.

Baseline procedures included a detailed history of past medical events, physical examination, and anthropometrics to assess nutritional status was assessed using weight-for-age and height-for-age by age- and sex-normalized z-scores as defined by World Health Organization (WHO) global standards.^15^ Malnutrition is defined by WHO/UNICEF as weight- and/or height-for-age lower than <2.0 z-scores for age and sex (Table 1).^14^ Other measurements at enrolment and months 18 and 30 milestone time points included percutaneous oxygen saturation (O_2_ saturation; Arcatron Oximeter model X004C, India), blood pressure, complete blood count, renal and hepatic function tests (Figure 1).

**TABLE 1.**
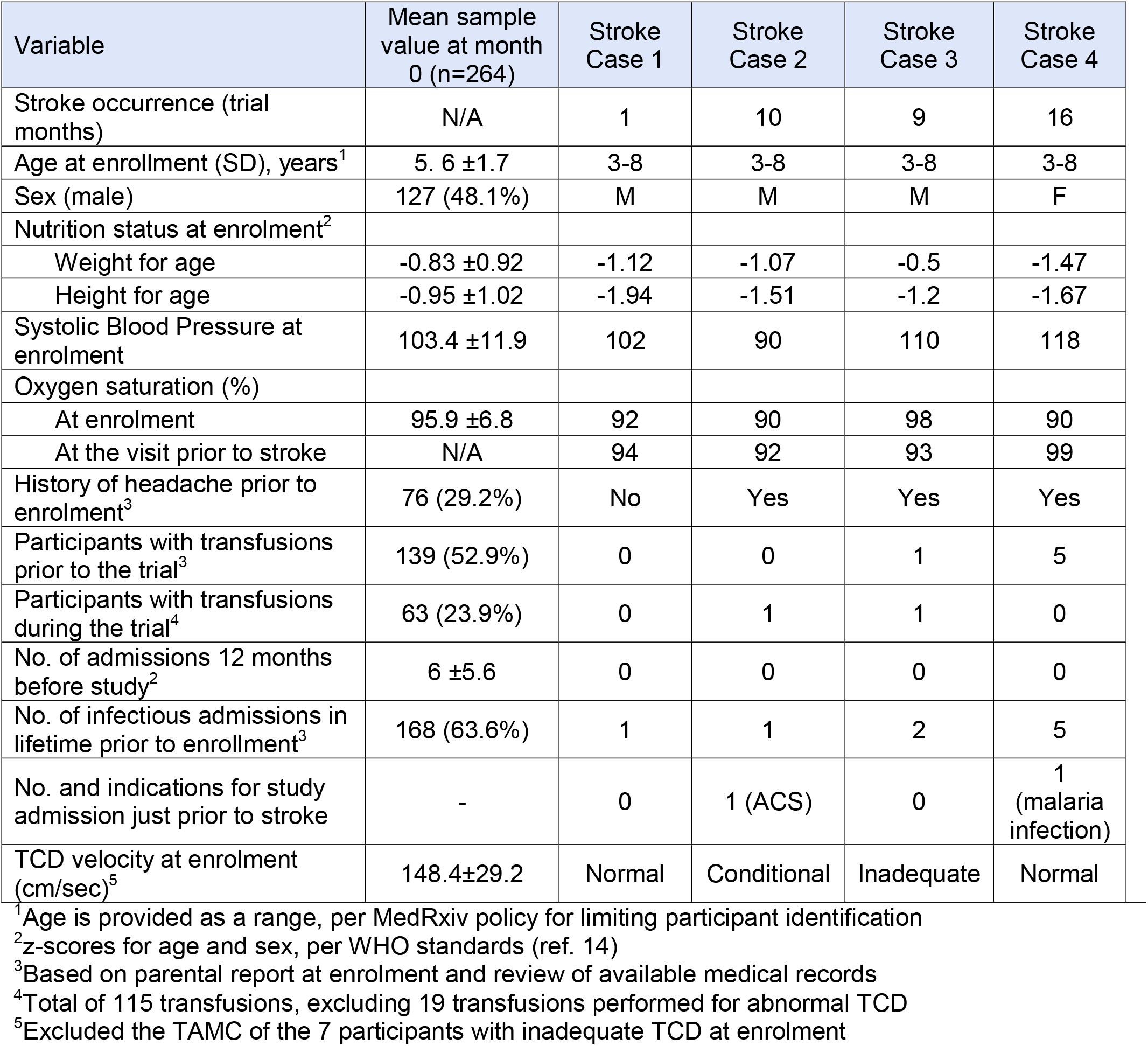
Trial demographic and other characteristics in the overall sample compared to the 4 participants who developed acute stroke.

| Variable | Mean sample value at month 0 (n=264) | Stroke Case 1 | Stroke Case 2 | Stroke Case 3 | Stroke Case 4 |
| --- | --- | --- | --- | --- | --- |
| Stroke occurrence (trial months) | N/A | 1 | 10 | 9 | 16 |
| Age at enrollment (SD), years<sup>1</sup> | 5.6 ±1.7 | 3-8 | 3-8 | 3-8 | 3-8 |
| Sex (male) | 127 (48.1%) | M | M | M | F |
| Nutrition status at enrolment<sup>2</sup> |  |  |  |  |  |
| Weight for age | -0.83 ±0.92 | -1.12 | -1.07 | -0.5 | -1.47 |
| Height for age | -0.95 ±1.02 | -1.94 | -1.51 | -1.2 | -1.67 |
| Systolic Blood Pressure at enrolment | 103.4 ±11.9 | 102 | 90 | 110 | 118 |
| Oxygen saturation (%) |  |  |  |  |  |
| At enrolment | 95.9 ±6.8 | 92 | 90 | 98 | 90 |
| At the visit prior to stroke | N/A | 94 | 92 | 93 | 99 |
| History of headache prior to enrolment<sup>3</sup> | 76 (29.2%) | No | Yes | Yes | Yes |
| Participants with transfusions prior to the trial<sup>3</sup> | 139 (52.9%) | 0 | 0 | 1 | 5 |
| Participants with transfusions during the trial<sup>4</sup> | 63 (23.9%) | 0 | 1 | 1 | 0 |
| No. of admissions 12 months before study<sup>2</sup> | 6 ±5.6 | 0 | 0 | 0 | 0 |
| No. of infectious admissions in lifetime prior to enrollment<sup>3</sup> | 168 (63.6%) | 1 | 1 | 2 | 5 |
| No. and indications for study admission just prior to stroke | - | 0 | 1 (ACS) | 0 | 1 (malaria infection) |
| TCD velocity at enrolment (cm/sec)<sup>5</sup> | 148.4±29.2 | Normal | Conditional | Inadequate | Normal |

<sup>1</sup>Age is provided as a range, per MedRxiv policy for limiting participant identification
<sup>2</sup>z-scores for age and sex, per WHO standards (ref. 14)
<sup>3</sup>Based on parental report at enrolment and review of available medical records
<sup>4</sup>Total of 115 transfusions, excluding 19 transfusions performed for abnormal TCD
<sup>5</sup>Excluded the TAMC of the 7 participants with inadequate TCD at enrolment

**Figure 1.**
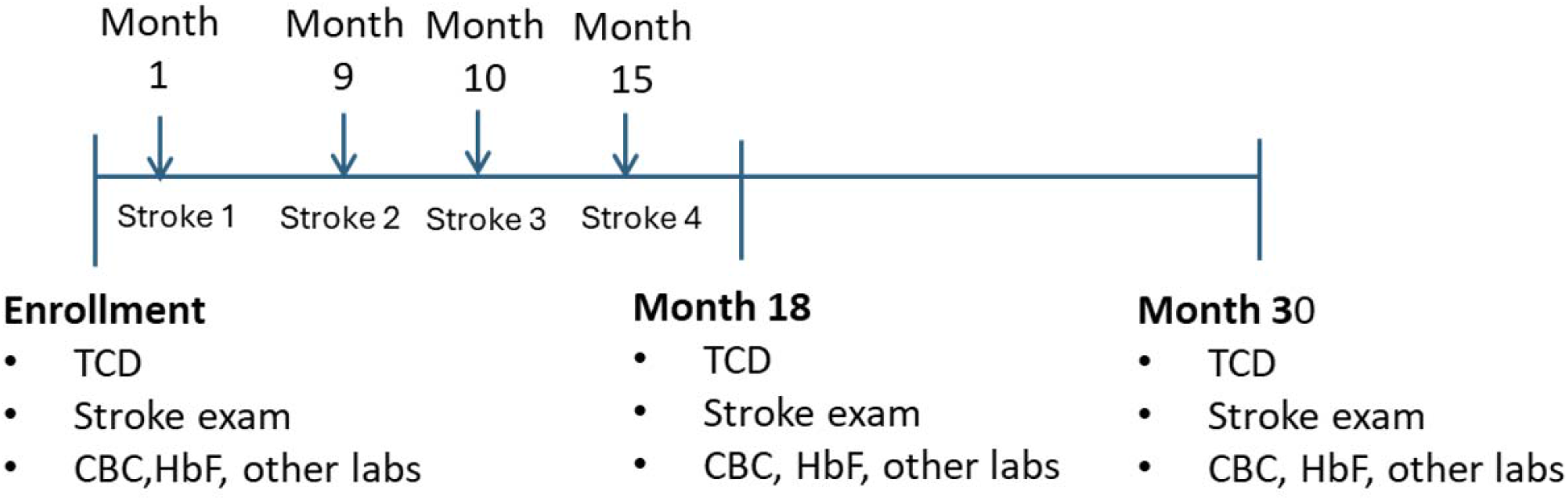
Trial timeline including incident strokes.

**Figure 2.**
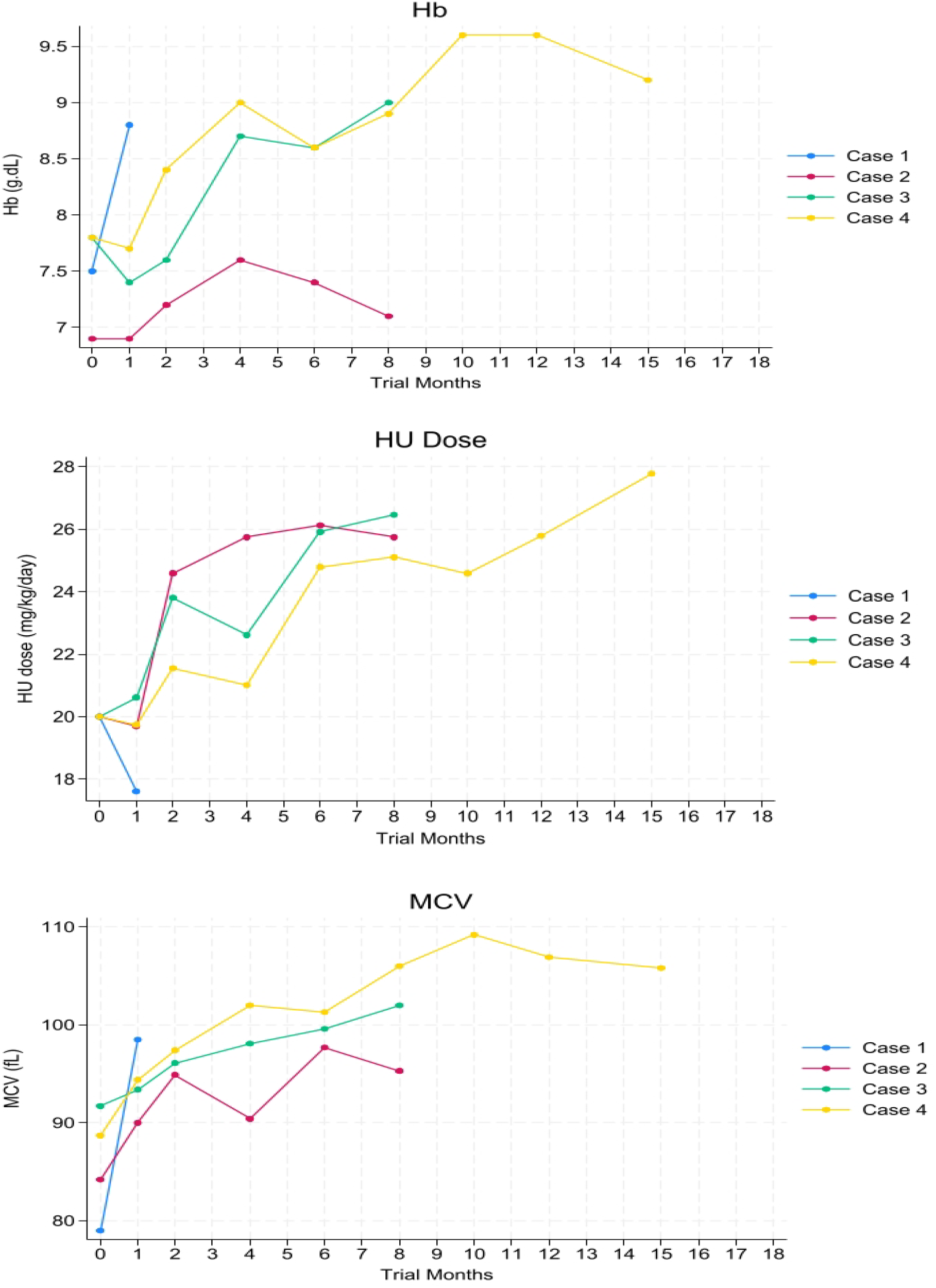
Trends of Hb, MCV and HU dose for the 4 cases of incident stroke.

Assessment at these three time points included TCD TAMV determination in 8 out of the 9 cerebral arteries (excluding the basilar artery) described in the STOP Trial using a portable non-imaging TCD machine (SonaraTek, Natus neurology, USA).^6^ Two co-authors (DM, MN), each were certified to perform TCDs at a U.S. pediatric academic medical center, provided TCD training, supervision and implementation of standard quality assurance measures for tester reliability to the four-study staff (two physicians, two nurses) who conducted trial TCD assessments. Trial TCD velocities were classified into categories using the STOP criteria: normal (<170□cm/sec), conditional (170-199□cm/sec), abnormal (≥200□cm/sec), or inadequate evaluation9. Participants with an abnormal TAMV were offered monthly blood transfusions, when available, to raise Hb level to 9-10g/dL. Participants with abnormal TCD had repeat TCD after one month and then every 6 months until TAMV improved or was declared persistently abnormal.

Upon completion of baseline trial assessments, HU therapy was initiated at 20±2.5mg/kg/day using scored tablets (Siklos, AddMedica (subsequently Theravia), France) for accurate weight-based dosing. Sufficient HU was provided to participant caregivers until the next scheduled trial visit. Escalation to maximum tolerated dose (MTD) by dose adjustment every 8 weeks unless a defined dose-limited hematological toxicity (DLT) occurred or the participant reached the MTD maximum of 30mg/kg daily.^20^ Escalation targets for hematologic values were an absolute neutrophil count (ANC) of 2.0-4.0 x10^9^ per liter and other standard hematologic, hepatic and renal criteria.^17,19^ We followed concurrent MHSCC guidelines for transfusion of Hb <5.0 or <6.0 if febrile.^18^

Following an initial 1-month study drug safety visit, regular trial visits assessed interim health, weight for accurate dosing, laboratory monitoring for drug safety, and assessment of adherence to daily trial drug scheduled at two-month intervals during the initial 12 months and subsequently at three-month intervals until trial completion.^18^ Caregivers were encouraged to adhere to daily HU use, which was monitored at study visits by assessing residual pill counts, laboratory-responsive blood count monitoring, especially ANC and red cell mean corpuscular volume (MCV). Serious adverse events (SAEs) were defined as death, incident stroke or prolonger hospitalization (>7 days).

### Data management and statistical analysis

Data were collected using electronic trial visit forms, with data entered and stored in the research database (REDCap™) for secure, web-based data management, hosted on a local server at Global Health Uganda (https://globalhealthuganda.org) with cloud back-up. Statistical analyses were conducted using STATA version 18. Descriptive statistics were used to summarize participant characteristics, including key demographic, clinical and laboratory factors, and TCD TAMV. Changes were analyzed using Student’s t-tests; statistical significance was defined as p <0.05.

## Results

Enrolment began in March 2021, with trial procedures completed in July 2024. The initial trial year coincided with the peak of COVID-19 pandemic in Uganda. A total 334 children were screened and 270 underwent preliminary enrolment. Five were subsequently excluded after confirmatory Hb electrophoresis failed to demonstrate SCA and one declined to continue. A total of 264 participants started on HU. Of these, 127 (48.1%) were male, mean age was 5.6±1.7 years, mean Hb 7.8±1.2 g/dL, mean HbF 11.9±8.1% and mean MCV 81.1±8.2fL (Tables 1 and 2). Initial mean TCD TAMV was 148.4± 29.2cm/second.

**TABLE 2.** Trial assessments of hematological parameters and hydroxyurea dose of the entire sample and the four participants with incident stroke.

| Variable | Mean value at month 0 (N=264) | Stroke Case 1 | Stroke Case 2 | Stroke Case 3 | Stroke Case 4 |
| --- | --- | --- | --- | --- | --- |
| Hemoglobin (g/dL) |  |  |  |  |  |
| At enrolment | 7.8±1.15 | 7.5 | 6.9 | 7.8 | 7.8 |
| At study visit prior to stroke | N/A | 8.8 | 7.1 | 9.0 | 9.2 |
| HbF (%) at enrolment<sup>1</sup> | 11.9±8.1 | 2.6 | 6.0 | 6.7 | 3.1 |
| HU dose at stroke (mg/kg/day) |  |  |  |  |  |
| At baseline | 20.17±0.91 | 18.25 | 19.84 | 18.92 | 20.55 |
| At visit prior to stroke | N/A | 17.6 | 25.7 | 26.5 | 27.8 |
| MCV (femtoliters) |  |  |  |  |  |
| At baseline | 81.1±8.24 | 79 | 84.2 | 91.7 | 88.718 |
| At visit prior to stroke | NA | 98.5 | 95.3 | 102 | 105.8 |
| No. DLTs on study<sup>2</sup> | N/A | 0 | 0 | 0 | 0 |
| ARC (x10<sup>9</sup>/L) |  |  |  |  |  |
| At enrolment | 0.35±0.11 | 0.49 | 0.45 | 0.37 | 0.43 |
| At visit prior to stroke | NA | 0.43 | 0.33 | 0.22 | 0.25 |
| WBC (x10<sup>6</sup>/L) |  |  |  |  |  |
| At enrolment | 13.8±4.2 | 19.35 | 22.06 | 14.11 | 15.54 |
| At visit prior to the stroke | NA | 15.26 | 16.09 | 17.04 | 16.09 |
| ANC (x10<sup>6</sup>/L) |  |  |  |  |  |
| At enrolment | 5.59±2.3 | 5.48 | 5.57 | 4.84 | 5.81 |
| At visit prior to stroke | NA | 7.08 | 7.63 | 11.87 | 9.42 |
| Platelets x10<sup>6</sup>/L |  |  |  |  |  |
| At enrolment | 443.7±153.0 | 452 | 541 | 336 | 552 |
| At visit prior to stroke | NA | 494 | 496 | 633 | 624 |

<sup>1</sup>Reached premature endpoint prior to re-testing at planned trial midpoint
<sup>2</sup>DLT: Asymptomatic dose-limiting toxicity
NA: Not applicable

HU therapy was initiated at mean daily dose of 20.2±0.9 mg/kg, and escalated to mean MTD dose at 25.9±2.7mg/kg.^17^ At trial completion, adverse events (AEs) totaled: 134 blood transfusions (21.2 per 100 participant years) (excluding transfusions provided for abnormal TCD), 429 medically treated pain crises (67.7 per 100 participant years) and 97 dose-limiting laboratory toxicities (15.8 per 100 participant years) (Table 2). Four participants had prematurely withdrawn or were lost-to-follow-up. SAEs were: 4 deaths from disease and/or infection-related complications without evidence of acute stroke. There were also 4 non-fatal acute strokes (Table 1), an incidence of 0.62 per 100 person years; having reached a premature trial endpoint, they were transitioned to the stroke program of the MHSSC. Additional SAEs included 9 prolonged admissions. HU-related AEs were asymptomatic dose-limiting toxicities; no HU-related SAEs occurred. One child underwent splenectomy for chronic hypersplenism which prevented HU therapy; she resumed study drug after an uneventful recovery. Baseline and milestone laboratory data demonstrated mean laboratory data, focused on HU-responsive hematological data (Table 2), including mean Hb increase from 7.8±1.2 to 8.8±1.5g/dL.

### TCD velocity and stroke risk

At enrolment, 43 participants (16.3%) had a conditional TCD and 15 (5.7%) had an abnormal TCD (Table 3.) At month 18 TCD, repeat TCD revealed a mean TAMV decreased with HU therapy by 17.4 cm/sec, from 148.4± 29.2 to 130.9±21.3cm/sec. At month 30, mean TAMV was 131.9±SD25.7; 15 (6.0%) had a conditional and 6 (2.4%) had an abnormal TCD.

**TABLE 3.**
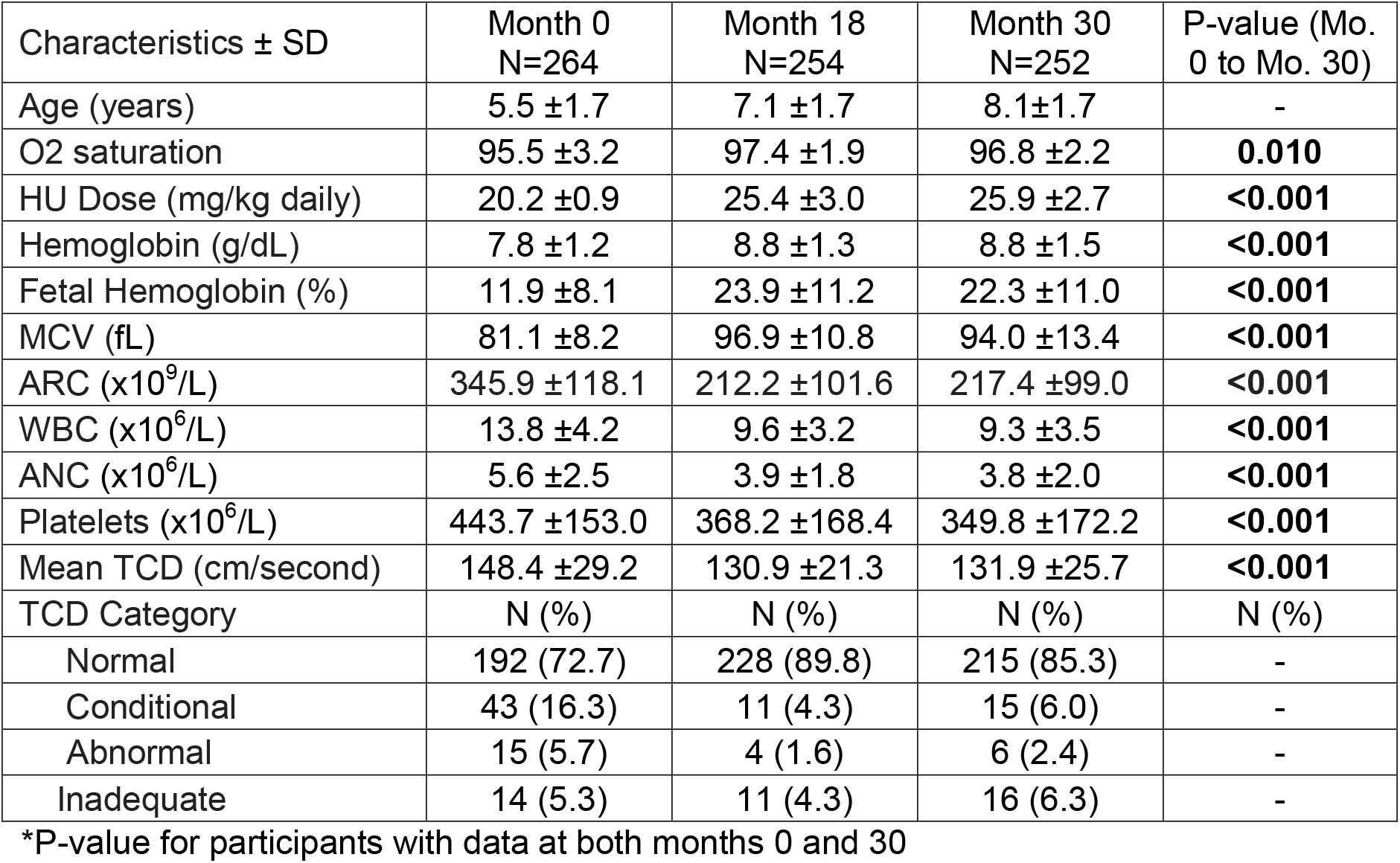
Clinical, laboratory and TCD TAMV for the entire sample, assessed at months 0, 18 and 30.

| Characteristics ± SD | Month 0<br>N=264 | Month 18<br>N=254 | Month 30<br>N=252 | P-value (Mo.<br>0 to Mo. 30) |
| --- | --- | --- | --- | --- |
| Age (years) | 5.5 ±1.7 | 7.1 ±1.7 | 8.1±1.7 | - |
| O2 saturation | 95.5 ±3.2 | 97.4 ±1.9 | 96.8 ±2.2 | **0.010** |
| HU Dose (mg/kg daily) | 20.2 ±0.9 | 25.4 ±3.0 | 25.9 ±2.7 | **<0.001** |
| Hemoglobin (g/dL) | 7.8 ±1.2 | 8.8 ±1.3 | 8.8 ±1.5 | **<0.001** |
| Fetal Hemoglobin (%) | 11.9 ±8.1 | 23.9 ±11.2 | 22.3 ±11.0 | **<0.001** |
| MCV (fL) | 81.1 ±8.2 | 96.9 ±10.8 | 94.0 ±13.4 | **<0.001** |
| ARC ($\times 10^9$/L) | 345.9 ±118.1 | 212.2 ±101.6 | 217.4 ±99.0 | **<0.001** |
| WBC ($\times 10^6$/L) | 13.8 ±4.2 | 9.6 ±3.2 | 9.3 ±3.5 | **<0.001** |
| ANC ($\times 10^6$/L) | 5.6 ±2.5 | 3.9 ±1.8 | 3.8 ±2.0 | **<0.001** |
| Platelets ($\times 10^6$/L) | 443.7 ±153.0 | 368.2 ±168.4 | 349.8 ±172.2 | **<0.001** |
| Mean TCD (cm/second) | 148.4 ±29.2 | 130.9 ±21.3 | 131.9 ±25.7 | **<0.001** |
| TCD Category | N (%) | N (%) | N (%) | N (%) |
| Normal | 192 (72.7) | 228 (89.8) | 215 (85.3) | - |
| Conditional | 43 (16.3) | 11 (4.3) | 15 (6.0) | - |
| Abnormal | 15 (5.7) | 4 (1.6) | 6 (2.4) | - |
| Inadequate | 14 (5.3) | 11 (4.3) | 16 (6.3) | - |

\*P-value for participants with data at both months 0 and 30

### Participants with incident stroke

The incident strokes occurred at trial months 1, 9, 10 and 15 months (mean 9.3 months) (Table 1). Those 4 participants had been enrolled from ages 3 to 8 years; 3 were male. One of the 4 had a history of five pre-trial episodes of severe anemia necessitating transfusion. Three had history of headaches, as did 29.2% of the cohort. At enrolment, 2 had low oxygen saturation of 90% (defined as <95%), none were malnourished per WHO global criteria at enrolment and none had hypertension (Table 1).^15,23^ Two of these 4 children had HbF of ≤3.1%, compared to the entire samples of 11.9±8.1 (*p*=0.035); (Table 2).

At the scheduled visit preceding the stroke, all were on hydroxyurea therapy, with mean daily dose of 24.4±4.6mg/kg (range 17.6 to 27.8 mg/kg). Two in this small sub-sample had low O2 saturation. Mean Hb was 8.5±1.0g/dL, similar to the mean from the entire cohort at a comparable study time. Repeat HbF was not planned until trial month 18. Apparent stroke precipitants were severe malaria infection, acute chest syndrome (ACS) and recent pain crisis; one was of uncertain cause, as detailed (Table 1). Brief case descriptions of participants with incident stroke are in Clinical Case Material.

At trial completion, 8 of the 252 (3.2%) of active participants who had a normal TCD at enrolment had developed a higher risk category by trial completion. Among these 8, mean enrolment age 5.6±1.8 years, 6 had developed a conditional TCD and 2 an abnormal TCD. Clinical and laboratory review identified only a modest HU-induced increased mean Hb at enrolment compared to completion: 7.6±1.1 versus 8.1±1.1g/dL (p=0.86) and HbF (13.3±8.5 versus 17.3±12.7 (p=0.35), despite escalated mean HU dose (27.6±1.8mg/kg) and increased mean MCV from 80.1±6.3 to 93.4±11.4, p=0.004). No other outlying values or patterns were observed (data not shown).

## Discussion

Here we report the findings for TCD and incident stroke from our Uganda-based our 30-month, single-arm open-label pediatric SCA HU trial with dose escalation to assess brain protection. Standard TCD screening was used as the primary stratification for identifying high stroke risk and to target intensified intervention. Reduced mean TCD velocity and the proportion of participants with elevated risk category were accomplished and concordant with increased cohort mean Hb level of 1g/dL.^24^ Rate of incident stroke was comparable to that reported in a recent pediatric HU stroke prevention trial in Nigeria.^7^ In contrast, none of our four participants with incident stroke had an abnormal TCD at enrolment. This finding suggests additional risk factors beyond an abnormal TCD may be important precipitants of acute stroke in the region.

At trial completion, a *post hoc* analysis was performed of the 4 participants with incident stroke and 8 with a worse TCD category compared to enrollment to identify additional stroke risk factors and precipitants beyond abnormal TCD.^2,21,23^ Two of the four with stroke had well-recognized precipitants of acute SCA- and/or infection, while the other two lacked an evident precipitating event. That incident strokes occurred within months 1-16 of the 30-month trial suggests potentially pre-existing risks, despite lack of an abnormal entry TCD. Historical, clinical and/or laboratory risks identified at enrolment included: 1) one child had a conditional TCD, signifying some degree of abnormal cerebral blood flow and one with an inadequate TCD. These events aligned with the increased risk associated with both conditional and inadequate TCD categories already identified in prior SCA studies, especially in children with no previous TCD assessment; 2) two children had very low HbF levels, well established as a disease marker of severity; 3) two had low O2 saturation, a known risk factor for SCA cerebrovascular events; and 4) one with prior recurrent episodes of severe anemia.^9,25,26^ Neither chronically very low Hb nor the presence of malnutrition or hypertension were present among the small number with incident stroke. Among the 8 participants with worse TCD category at trial completion, modest HU-induced Hb and HbF responses may have adversely affected their TCD category.

A recent stroke prevention trial in Tanzania with intensive HU dosing similar to our own approach reported no incident strokes over a 12-month period.^8^ In contrast, a phase 3 trial in Nigeria reported incident strokes at twice the rate we report here, but enrolment was limited to children with abnormal TCD.^7^ A prospective observational TCD study in the region reported similar predictors for stroke, including low Hb, low O_2_ saturation and young age.^9^ Chronic risk factors such as hypoxia or hypertension may also increase the risk of vasculopathy and subsequent stroke.^26,27^ Additional risks may arise from beta globin haplotypes in the region and alpha thalassemia trait, as well as acute and chronic infections and other potential region-related risks in this vulnerable population.^14,15,28,29^ Taken together, these findings suggest that the participants in our Ugandan trial with acute stroke had risk factors which dose-intensive HU therapy could not fully overcome.

In the U.S.-based SWiTCH Trial, dose-escalated HU therapy did not fully prevent recurrent stroke in children with severe cerebral vasculopathy identified by magnetic resonance imaging (MRI), nor as primary stroke prevention in the TWiTCH Trial in those with abnormal TCD.^30,31^ A randomly selected subset of 89 (34%) participants within our trial, primarily ages 5 years and older, underwent MRI/MRA imaging on a 1.5 tesla scanner (without sedation) at enrolment and month 30 trial completion (to be reported separately). Preliminary analyses suggest that over half of the sub-sample imaged had silent cerebral infarcts and several had vascular stenosis. As imaging is not standard of care in the region, only a subset was imaged, and every participant was on trial HU, the protocol required intervention for abnormal TCD but not for MR-imaged abnormalities. Upon review for correlation with incident stroke TCD, 3 of the 4 participants with stroke had undergone MRI imaging at enrolment. All 3 had sickle cerebral arteriopathy, despite not having had abnormal TCD. Having reached a premature endpoint, those with stroke were not re-imaged. Among the 8 children with worsened TCD category, only 2 had been MRI-imaged: both had non-progressive clinically silent cerebral infarcts, among which one had an arterial stenosis at enrolment.

Despite substantial reduction in mean TCD TAMV, a unifying risk among trial participants was that SCA diagnosis and care early in life, including continuous HU therapy, were not uniformly available for potential protection from early onset SCA cerebrovascular injury.^32^ A U.S. pediatric observational study conducted during the pre-HU era reported the same incident stroke rate for SCA as reported here.^33^ This equivalence suggests that BRAIN SAFE-II participants had pre-existing and/or ongoing stroke risk, despite intensive HU therapy. The older U.S. study had also reported higher stroke risk with silent cerebral infarcts (SCI), consistent with our finding from a prior Ugandan observational study of >60% SCI prevalence.^34,35^

### Limitations

Our study had a small number of participants with stroke from which to identify risk factors. Nonetheless, trial use of the PedNIHSS as a sensitive tool for prior stroke identification likely enhanced the focus on primary stroke prevention.^11,20,24^ Our study assessed a larger number of children over a longer period of time (636 person years) than most other studies in the region.^11,20,24^ Strokes occurred prior to the scheduled repeat TCD, precluding assessment of HU effect for those participants. We did not investigate potential cardiac or pulmonary etiologies of low O2 saturation. Funding was insufficient for MRI/MRA imaging for all trial participants.

In conclusion, abnormal TCD velocity alone appeared to be insufficient to fully identify children who developed an acute stroke during the Ugandan trial despite intensive HU therapy. Our findings also suggest a high degree of early cerebrovascular risk among these children with SCA. Known factors for identifying elevated stroke risk include elevated or inadequate TCD velocity, severe anemia, low HbF, low O2 saturation, hypertension, and history of headache.^26^ Recognized SCA-related stroke triggers include complications such as severe infection, ACS and, to a lesser extent, painful crises. Additional studied are needed to further understand stroke triggers and other cerebrovascular adversity in SSA and the role of TCD risk stratification in SSA. The last point was specifically identified in a recent meta-analysis as a key research need within low- and middle-income countries.^36^

Nonetheless, in addition to excellent – if imperfect – stroke protection through HU therapy, our findings and the contrast with reports from higher-resource countries on substantial stroke prevention in children with SCA suggest that early and continuous HU therapy be initiated to limit or prevent cerebrovascular injury.^14^ HU therapy may also benefit children in the region by providing additional intensive anti-infectious prophylaxis to reduce potential stroke precipitants of severe malaria and other serious infections.^37^ Further, children identified at higher stroke risk despite HU therapy, such as with persistent abnormal TCD, low levels of Hb, HbF and O2 saturation despite intensive HU therapy may need more aggressive stroke prevention, up to and including hematopoietic stem cell transplantation or gene therapy.^38^

## Data Availability

All aggregated, de-identified data produced in the present study are available upon reasonable request to the authors

## Abbreviation Complete Term

TCD: Transcranial doppler ultrasound
SCA: Sickle cell anemia
MTD: Maximum tolerated dose
HbF: Fetal hemoglobin
SSA: Sub-Saharan Africa
MHSCC: Mulago Hospital Sickle Cell Clinic
Hb: Hemoglobin
TAMV: Time-averaged maximum mean velocity
HU: Hydroxyurea
IRB: Institutional review boards
PedNIHSS: Pediatric NIH stroke scale
O2: saturation Percutaneous oxygen saturation
ANC: Absolute neutrophil count
MCV: Red cell mean corpuscular volume
AE or SAE: Adverse events or severe AEs
ACS: Acute chest syndrome
MRI: Magnetic resonance imaging

## Acknowledgements

This trial is registered as NCT04750707. The work was supported by grants the National Institutes of Health: Eunice Kennedy Shriver National Institute of Child Health and Human Development R01HD096559 (Idro, Green) and the Fogarty International Center D43TW010928 (John, Idro). Study drug (Siklos) was donated (Theravia, formerly AddMedica, France). The TOVA Company provided the instrument used at a discounted rate for this research.

The funders, donors or sponsors had no role in designing, gathering, analyzing, or interpreting the data. The content of this publication does not necessarily reflect the views or policies of the Department of Health and Human Services, nor does mention of trade names, commercial products, or organizations imply endorsement by the US government.

We gratefully acknowledge the support of the outstanding study team and from Global Health Uganda. We also thank the Pediatric staff of the MHSCC, who served as key clinical links to the trial and the Data Safety and Monitoring Committee. Most importantly, we thank the many families who supported their child’s trial participation.

## Clinical Case Material

Case 1 was a boy 3-8 years of age who developed acute neurological symptoms of facial asymmetry after one month on study. Symptoms were preceded by a vaso-occlusive crisis 2 weeks earlier. There was no history of fever or other signs or symptoms of an acute infectious illness. Oxygen saturation was 94% on room air; hemoglobin was 8.8g/dL.

Case 2 was a boy 3-8 years of age who developed cough, fever and vomiting for 4 days associated with difficulty breathing. He was febrile with tachypnea and tachycardia, with oxygen saturation of 91% on room air and had bilateral crackles in his lung fields. Hemoglobin was 6.5g/dL. Malaria smear was negative. He was hospitalized for 10 days for acute chest syndrome with blood transfusion, oxygen therapy, intravenous antibiotics and oral analgesics. He developed right-sided weakness on day 3 of the admission. Of note, his Hb had dropped by 0.5g/dL between months 4 and 8 despite increase in HU dose with relative increase in MCV through month 8 without a palpable spleen.

Case 3 was a boy 3-8 years of age who developed sudden onset of unilateral weakness while playing at home, followed by a generalized tonic-clonic seizure. There had been no symptoms suggestive of an infectious etiology. Hb was 7.8g/dL. The blood slide for malaria showed no parasites. Soon thereafter he developed aspiration pneumonia.

Case 4 was a girl 3-8 years of age with fever and cough for 1 day and associated seizures with an episode of loss of consciousness who also developed aphasia and right-sided weakness. Hb was 9.2g/dL. There was no prior history of illness. Evaluation was notable for malaria parasitemia on blood smear, indicating and acute malaria infection.

